# The Hidden Gap in Infant HIV Diagnosis: A Call for Complementary Diagnostics in the Point-of-Care and Prophylaxis Era

**DOI:** 10.1101/2025.07.28.25331911

**Authors:** Grace Esther Kushemererwa, Oreen Kemigisha, Christine Namulindwa, Susan Nabadda, Isaac Ssewanyana

## Abstract

The remarkable success of Prevention of Mother-to-Child Transmission (PMTCT) programs has significantly lowered infant HIV prevalence. However, this success, along with widespread maternal and infant prophylaxis and the increasing reliance on Point-of-Care (POC) diagnostics, has inadvertently created a new diagnostic challenge. Our research quantifies a critical, previously overlooked gap in HIV detection among infants born to HIV-positive mothers. We observed that a substantial proportion (over 18%) of HIV-infected infants now present with extremely low viral loads (Ct values above 31), which is likely due to the suppressive effects of prophylaxis. Importantly, external quality assessment studies reveal that most current POC platforms in use cannot be single-handedly relied on to detect HIV at these low concentrations. This means that sole reliance on POC testing risks leaving a significant cohort of approximately 18% of HIV-infected infants undiagnosed for prolonged periods, leading to severe clinical outcomes and undermining PMTCT gains. We emphasize the essential role of highly sensitive conventional PCR platforms, capable of accurately interpreting very low positive results, and advocate for a complementary diagnostic strategy where conventional PCR serves as a confirmatory test for all HIV-exposed infants. This article highlights the urgent need for a revised diagnostic approach to optimize PMTCT strategies and ensure timely care for every HIV-exposed infant.

## Introduction

Early and accurate diagnosis of Human Immunodeficiency Virus (HIV) infection in infants born to HIV-positive mothers is essential for decreasing morbidity and mortality [1]. Untreated HIV-infected infants experienceface high death rates, with estimates showing about 30% mortality by the age of one and 50% by the age of two [2]. Timely initiation of Antiretroviral Therapy (ART) profoundly improves clinical outcomes, informs crucial infant feeding decisions, and facilitates the identification of HIV-exposed but uninfected infants for ongoing follow-up and preventive interventions [3].

Worldwide efforts, particularly through Prevention of Mother-to-Child Transmission (PMTCT) programs, have shown great success, which has resulted in a significant decrease in infant HIV prevalence [4]. This achievement is a result of the widespread implementation of maternal and infant antiretroviral prophylaxis. Although this preventative treatment has been highly effective, it has also created an unexpected diagnostic challenge: a growing proportion of HIV-infected infants now present with significantly suppressed viral loads, complicating detection with standard Point of Care Tests. This article measures the true extent of the under-detection problem and suggests a vital change to how HIV infant diagnosis is currently made using the Point of Care Tests.

Although POC comes with a lot of advantages, the sensitivities of the POC machines in use is low compared to the convention Laboratory based platforms. Some of these POC can only detect HIV in specimen whose viral load (VL) is above 500 copies/ml. It has been observed that POC tests may fail to detect the virus in samples with very low viral concentrations, characterized by high cycle threshold (Ct) values (e.g., Ct value greater than 30). This raises significant concerns that POC tests might miss a substantial number of true HIV infections in infants, requiring the urgent need for improvements in the HIV testing algorithm when using POC platforms.

## Methods

### Study Design

Our study was a retrospective review of Early Infant Diagnosis (EID) data. The study was conducted at the Central Public Health Laboratories (CPHL), Ministry of Health Uganda. All data were sourced directly from the CPHL Laboratory Information Management System (LIMS).

We reviewed all EID data tested from January 2023 to June 2023. For all infant samples tested for HIV EID, the following specific data points were extracted:

- Sample identifier
- Health facility name
- Date of sample collection
- Age of the infant at sample collection
- Number of PCR tests performed (if applicable)
- Date of sample reception at CPHL
- Date of testing
- Test result (confirmed HIV-infected or not)
- Quantitative PCR cycle threshold (Ct) values for all confirmed HIV-infected infant samples.

Data extraction and compilation were performed by trained personnel to ensure accuracy and completeness. Regular data quality checks were performed throughout the process to identify and rectify any inconsistencies or missing information, thereby ensuring the reliability of our dataset.

The extracted data was analyzed using Microsoft Excel Version 1808.

## Result

Out of 49,529 EID tests reviewed, 669 were confirmed as HIV positive. Further analysis using the Cobas HIV qualitative test revealed the following distribution of Ct values among the positive results:

1. 126 (18.8%) exhibited a Ct value exceeding 31. These cases represent extremely low viral loads and are most unlikely to be detected by many of the current POC platforms. This is highlighted by a red traffic light symbol (Figure. 2).
2. 55 (7.7%) demonstrated a Ct value ranging from 30 to 31. For these cases, there is approximately a 50% chance of detection by existing POC platforms. This category is marked with a yellow traffic light symbol, signifying some issues and medium criticality (Figure. 2).
3. 491 presented a Ct value below 30. These indicate higher viral loads, and all these cases are expected to be detected by existing POC platforms. This category is indicated by a green traffic light symbol, representing no issues and very desirable detection. (Figure 1)

**Figure 1.**
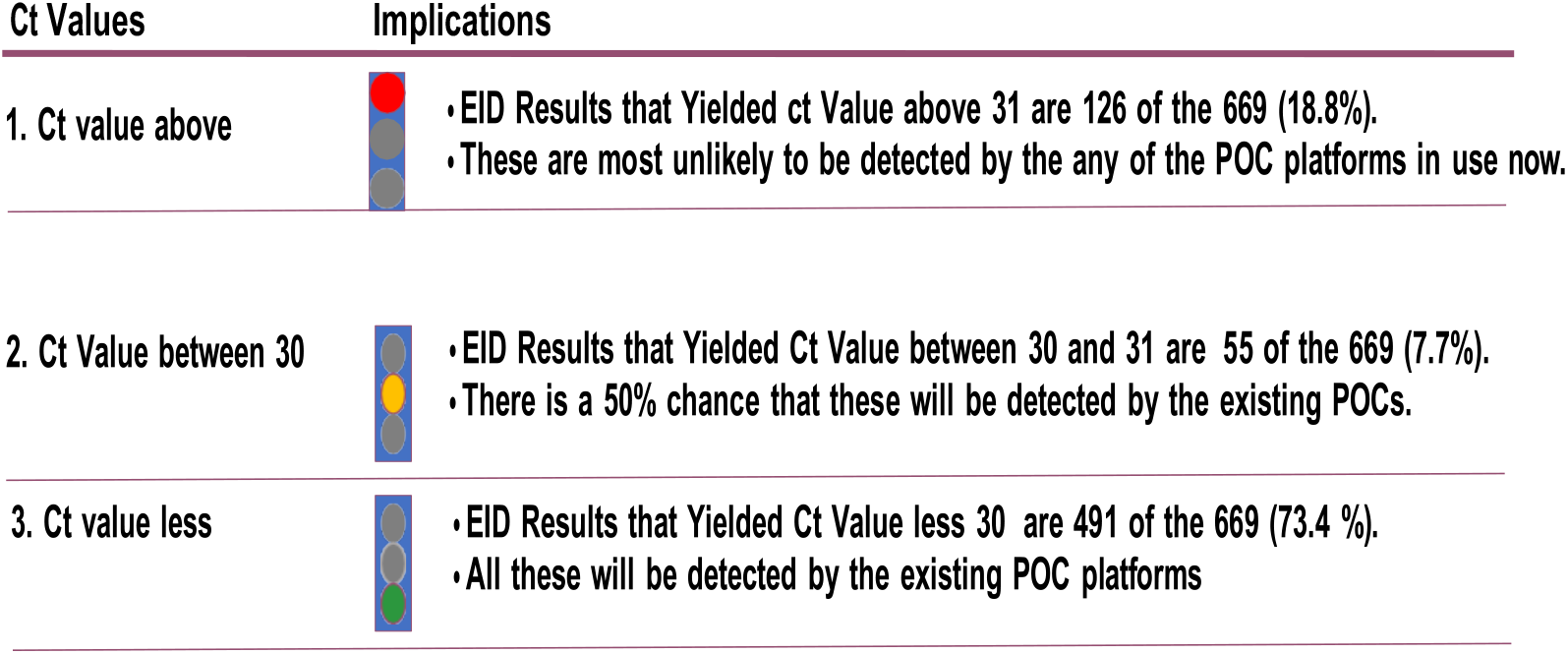
Distribution of ct values among positive results and likely hood of detection by POC Platform

This observation clearly indicates that while maternal and infant prophylaxis is effective in reducing transmission, it also sufficiently suppresses viral replication in a group of infected infants to levels near or below the limits of detection for many Points of Care platforms.

## Discussion

The implications of these findings are profound for global PMTCT strategies. If POC diagnostic tools are deployed as the sole testing platforms, a significant group of approximately 18% of HIV-infected infants could go undiagnosed for long periods. Such delays in diagnosis translate directly into delayed ART initiation, escalating the risk of severe opportunistic infections, irreversible immune compromise, and ultimately, preventable mortality. This diagnostic vulnerability poses a significant threat to achieving the global goal of an AIDS-free generation.

Our research emphasizes the vital role of highly sensitive conventional PCR platforms. These laboratory-based systems have significantly lower limits of detection and are very sensitive to accurately detect and interpret very low positive results, including those with high Ct values. This inbuilt sensitivity makes them critical for identifying the “hidden” infections in infants exposed to prophylaxis.

Therefore, we advocate for a necessary shift towards a complementary diagnostic strategy. In this new framework, highly sensitive conventional PCR platforms would serve as a mandatory confirmatory test for every infant born to an HIV-positive mother, especially when initial POC results are negative or where there is a strong clinical suspicion of infection despite a negative POC result. This dual-platform approach ensures that even infants with extremely low viral loads due to prophylaxis are accurately identified, enabling early ART initiation and comprehensive care.

## Conclusion

Our evidence strongly point to a significant detection gap with current POC diagnostics and makes a strong case for the indispensable role of highly sensitive conventional PCR platforms in a complementary testing strategy. Addressing this hidden gap is crucial for optimizing existing PMTCT strategies, ensuring that every HIV-exposed infant benefits from the full spectrum of available care, and ultimately achieving the aspirational goal of an AIDS-free generation. We believe these findings warrant urgent discussion and action among researchers, clinicians, and policymakers to refine diagnostic algorithms and safeguard the health of the most vulnerable population.

## Data Availability

HIV Test dataa

